# First proof of the capability of wastewater surveillance for COVID-19 in India through detection of genetic material of SARS-CoV-2

**DOI:** 10.1101/2020.06.16.20133215

**Authors:** Manish Kumar, Arbind Kumar Patel, Anil V Shah, Janvi Raval, Neha Rajpara, Madhvi Joshi, Chaitanya G Joshi

## Abstract

we made the first ever successful effort from India to detect the genetic material of SARS-CoV-2 viruses to understand the capability and application of WBE surveillance in India. Sampling was carried out on 8 and 27 May, 2020 from Old Pirana Waste Water Treatment Plant (WWTP) at Ahmedabad, Gujarat with 106 million liters per day (MLD) capacity receiving effluent of Civil Hospital treating COVID-19 patient. All three i.e. ORF1ab, N and S genes of SARS-CoV-2 were discerned in the influents with no gene spotted in the effluent collected on 8 and 27 May 2020. Temporal difference between 8 and 27 May 2020 samples was of 10x in gene copy loading with corresponding change of 2x in the number active COVID-19 patient in the city. Number of gene copies was found comparable to that reported in the untreated wastewaters of Australia, China and Turkey and lower than that of the USA, France and Spain. This study, being the first from India and probably among the first ten reports in the world of gene detection of SARS-CoV-2 in the environmental samples, aims to assist concerned authorities and policymakers to formulate and/or upgrade the COVID-19 surveillance to have explicit picture of phase of the pandemic. While infectious SARS-CoV-2 has yet to be identified in the aquatic environment, the virus potentially enters the wastewater stream from patient excretions and thus can be a great tool for pandemic monitoring.

**HIGHLIGHTS:**

□ First ever report of the presence of gene of SARS-CoV-2 in the wastewater in India.
□ C_T_ value is explicitly indicative of the increase of COVID-19 patient in the vicinity.
□ All three i.e. ORF1ab, N and S genes of SARS-CoV-2 were discerned in the influents.
□ None of three genes were spotted in the effluent collected on 8 and 27 May 2020.
□ Wastewater surveillance conclusively specified temporal difference in COVID-19 load.
□ Temporal difference was 10x and 2x in gene copies and COVID-19 patient, respectively.

## Introduction

The current ongoing global Coronavirus disease (COVID-19) pandemic, caused by the infection of severe acute respiratory syndrome coronavirus 2 (SARS-CoV-2), has spread to 216 countries and territories, with 7.7 million of the confirmed cases and more than 425,000 deaths worldwide, as of June 12, 2020 (WHO, 2020). The active replication of infectious SARS-CoV-2 particles in enterocytes of human intestine due to expression of ACE2 receptor causes shedding of virus in the feces (Lamers et al. 2020; Qi et al., 2020). The clinically reported symptoms in COVID-19 patients majorly include cough, difficulty in breathing, fever and diarrhoea (Gao et al., 2020; Kumar et al., 2020). However, during a previous investigation study of COVID-19 patients, higher proportion of the patients (48.1%) were reportedly detected the SARS-CoV-2 RNA in their feces samples than that of the patients exhibiting the gastrointestinal symptoms (17%) such as diarrhoea (Cheung et al., 2020). This results suggested that large number of asymptomatic individuals along with symptomatic patients discharge viruses shredded in the feces, which ultimately reaches to sewage treatment plants (Haramoto et al., 2020). These viruses can be shed in feces for several days, even after the patient stops exhibiting all the respiratory symptoms (Wu et al., 2020). As per a study reported by Zheng et al., 2020, the SARS-CoV-2 RNA in feces can be detected for median duration of 22 days. Though the residence time of SARS-CoV-2 virus has not been studied well, the evident studies reporting detection of SARS-CoV-2 RNA suggests possibility of detection of SARS-CoV-2 in the wastewater.

Wastewater based epidemiology is a promising approach to understand the status of the disease outbreak in a certain catchment by monitoring the viral load in the wastewater, as it contains the excretion from both symptomatic and asymptomatic individuals (Xagoraraki and O’Brien, 2020; Choi et al. 2018; Yang et al., 2015). WBE had been an effective tool during past outbreak of other enteric viruses, such as poliovirus, hepatitis A and norovirus (Hellmér et al., 2014; Asghar et al., 2014), it can be used as an early warning tool for the disease outbreak in a community and used to inform the efficacy of the current public health interventions (Ahmed et al., 2020). WBE data can help to estimate actual infected population due to the virus, as it covers asymptomatic and pre-symptomatic patients too, which may be underestimated by clinical surveillance (Tang et al., 2020; Wölfel et al., 2020a; Zhang et al., 2020).

Recently, SARS-CoV-2 RNA detection in wastewater has been reported in Australia, China, France, Israel, Italy, Japan, Netherlands, Spain and US (Ahmed et al., 2020; Bar- or et al., 2020; Haramoto et al., 2020; La Rosa et al., 2020; Medema et al., 2020; Nemudryi et al., 2020; Randazzo et al., 2020; Rimoldi et al., 2020; Wu et al., 2020; Wurtzer et al., 2020). According to some of these studies, after the number of confirmed cases reached to 1-100 cases per million population, SARS-CoV-2 RNA was detected in wastewater (Ahmed et al., 2020; Bar-or et al., 2020; Medema et al., 2020; Nemudryi et al., 2020; Wu et al., 2020; Wurtzer et al., 2020). To date, there is no single study reporting detection of SARS-CoV-2 in wastewater in India. As of June 12, 2020, the number of confirmed cases in India was 223 cases per million population. The first case of COVID-19 in India was reported on January 30, 2020 and the number of confirmed cases has reached to more than 300,000 as of June 12, 2020 (Ministry of Health and Family Welfare, India). The state of Gujarat has reported >22,500 confirmed cases of COVID-19, as of June 12, 2020, with >12,000 confirmed cases in Ahmedabad city. (Ministry of Health and Family Welfare, India).

Under these considerations, we made the first ever successful effort from India to detect the genetic material of SARS-CoV-2 viruses to understand the capability and application of WBE surveillance in India. We also analysed the temporal variation in genetic material loadings in the same wastewater treatment plant during lockdown period in India. Finally, we evaluated the effect of traditional treatment systems on SARS-CoV-2 genetic material. This study, being among the first ten reports in the world of gene detection of SARS-CoV-2 in the environmental samples, aims to assist concerned authorities and policymakers to formulate and/or upgrade the COVID-19 surveillance to have explicit picture of phase of the pandemic.

## Material and Methods

### Sampling

The wastewater samples were collected on 8 and 27 May, 2020 from Old Pirana Waste Water Treatment Plant (WWTP) at Ahmedabad, Gujarat which is the biggest wastewater treatment plant in Asia and has the capacity of 106 million liters per day (MLD). The WWTP is equipped with Upflow Anaerobic Sludge Blanket (UASB) as an advanced process to treat the wastewater. Specified design parameters for this WWTP is to produce a treated water with pH, biological oxygen demand (BOD), suspended solids (SS), and chemical oxygen demand of 7-8.5, > 20 mg.L-^1^, >30 mg.L and >100 mg.L^-1^ respectively. The plant is divided into three streams separated by three inlet chambers of 7.5 m x 5 m in size and the liquid depth of 2.5 m. Total of six grit chambers of 10.2 m × 10.2 m in size and water depth of 1.0m are provided (two in each stream) to remove gravel, sand and other settleable solids. Six Primary Clarifier (two in each stream) of 39.5 m in diameter and 3.2 m in depth are provided for providing a hydraulic retention time (HRT) of 2.5 hours for 60% and 30% reductions in SS and BOD, respectively. Six aeration tanks, each of 26.6 m x 60 m X 4.7 m in size are used for the secondary treatment process (biological treatment) which provides 5 hrs of hydraulic retention time. The corresponding secondary clarifiers are 43 m in diameter, 3.5 m in liquid depth with hydraulic retention time of 2.5 hours. The activated sludge is separated from the wastewater in this tank and settles down. The sludge from both the primary and secondary clarifiers are collected in respective sludge pits and pumped for sludge thickening and then further to sludge digestion tank for anaerobic digestion. Each stream has one sludge thickening unit (25 m diameter and liquid depth of 3.5 m) and two anaerobic digestion units (28 m diameter, 8 m liquid depth and retention time of 20 days).

Sampling location for this study was selected based on the fact that 106 MLD Pirana WWTP receives the sewage waste of government civil hospital treating COVID-19 patient. Wastewater samples (the influents, and the final effluents after UASB and aeration pond) were collected on 8 and 27 May 2020 to understand the temporal variation and the effect of wastewater treatment of SARS-CoV-2 RNA. In-situ water quality parameters (pH, temperature, electrical conductivity; EC, dissolved oxygen; DO, oxidation-reduction potential; ORP, and total dissolved solids) of the influent and effluent were noted using YSI Multiparameter probe prior to the sampling. Composite sample was made from three samples simultaneously taken in each location. Samples taken on 8 May were brought in the ice-box and refrigerated at 4 °C till 27 May when next batch of samples were brought to the laboratory and analysed on the same day. Sterile bottles were used for sampling and blanks in the same bottles were analysed to determine if there was any contamination during the transport. To ensure accuracy and precision, duplicated analyses of the samples was also performed. Several blanks were prepared and run to check the cross-contamination, and sensitivity of protocol, extraction and instrumentation. All analyses were done in Indian Council of Medical Research (ICMR), New Delhi approved facility of Gujarat Biotechnology Research Centre (GBRC).

### Method for the extraction of viral RNA from sewage samples

Viral RNAs were isolated from Sewage samples using following steps: Precipitation of viral particle; Viral RNA isolation and quality checking of RNA; RT-PCR analysis of viral RNA for the presence of SARS-CoV-2.

### Precipitation of viral particle

The sewage samples (50 mL) were centrifuged at 4500×g (Thermo Scientific) for 30 min followed by filtration of supernatant using 0.22 micron filters (Himedia). Further each sewage filtrate was concentrated using two methods: 1) using 96 well filter plate and 2) poly ethylene glycol (PEG) method. For the first method filtrate was concentrated using the 96 well filter plate (AcroPrep™ Advance 350 10K Omega™; Pall Corporation) and concentrate was further used as sample for RNA isolation. For the second method, PEG 9000 (80 g/L) and NaCl (17.5 g/L) was mixed in 25 ml filtrate and this was incubated at 17°C, 100 rpm for overnight. Next day the mixture was centrifuged at 13000×g (Kubota Corporation) for 90 mins. After centrifugation, supernatant was discarded and pellet was resuspended in 300µL RNase free water. This was further used as a sample for RNA isolation.

### RNA isolation

RNA isolation was carried out using commercially available kit (NucleoSpin^®^ RNA Virus, Macherey-Nagel GmbH & Co. KG, Germany). Concentrated viral particles were mixed with 10 µL MS2 phage (internal control provided by TaqPath™ Covid-19 RT-PCR Kit), 20 µL Proteinase K (20mg/mL) solution and 600 µL of RAV1 buffer containing carrier RNA. Further steps were carried out as instructed in product manual (Macherey-Nagel GmbH & Co. KG). RNA concentrations were analysed by Qubit 4 Fluorometer (Invitrogen).

### Real time PCR for the detection of SARS-CoV-2

RNAs were analysed for the detection of SARS-CoV-2 by RT-PCR using TaqPath™ Covid-19 RT-PCR Kit (Applied Biosystems). Procedures were carried out as described in product manual and interpretations of results was analysed as instructed in manual.

### Semiquantitative method for SARS-CoV-2 Gene detection

Although there is no direct correlation of the Ct value to copy numbers as the kit used for the detection is absent present assay but considering 500 copies of SARS-CoV-2 genes taken as positive control with Ct of average 26 for all the three genes i.e. ORF1ab, N and S, the same was extrapolated to compare it with sample Ct values and derive approximate copies of genes in the waste water sample. The amount of RNA used as template was multiplied with the enrichment factor to derive copy numbers for each waste water sample.

## Results and discussions

We examined three genes of SARS-CoV-2, ORF1ab, N protein genes and S protein genes, from the influent samples and the final effluent after USAB treatment and aeration pond, both from WWTP Pirana, on May 8 and May 27. MS2 phage was spiked in the samples as a inhibition control, and CT values were 22.46, 22.35 in the influents on May 8 and May 27, respectively, and 22.4 and 22.2 in the final effluents on May 8 and May 27, respectively. This indicates that there was no significant difference in inhibitory effects by wastewater matrix on RT-PCR performance among these samples. The positive control sample had CT values of the three SARS-CoV-2 genes ranging 27.92 to 29.52, while the SARS-CoV-2 genes were not detected from the negative control sample.

The results showed that the WWTP samples on both May 8 and May 27 were positive with all of ORF1ab, N protein genes and S protein genes, which were examined as SARS-CoV-2 genes, with the maximum concentration of 2.419×10^8^ copies/L (Table 1). To the best of our knowledge, this is the detection of SARS-CoV-2 genes in wastewater samples from India. ORF1ab assay and N protein assay have been used for SARS-CoV-2 RNA detection by RT-PCR from raw wastewater and river water samples in Milano, Italy (Rimoldi et al 2020). S protein gene has also been used for raw wastewater analysis in Italy (La Rosa et al 2020).

**Table 1.** Amplification cycles (C_T_) of raw wastewater and final effluents along with number of total case reported positive and discharged since 17^th^ March 2020 in Ahmedabad (AMD) and India (IND).

|  |  | ORF1ab | N protein gene | S protein gene | MS2 |
| --- | --- | --- | --- | --- | --- |
| Raw wastewater | 8-May | 35.52 | 35.39 | 39.56 | 22.46 |
|  | 27-May | 32.65 | 34.18 | 34.83 | 22.35 |
| Final effluent | 8-May | - | - | - | 22.40 |
|  | 27-May | - | - | - | 22.20 |
|  |  | Confirmed case<br>AMD | IND | Discharged/Recovered case<br>AMD | IND |
| *COVID-19 case<br>Since 17 March<br>2020 | 7-May | 4912 | 56352 | 985 | 16776 |
|  | 26-May | 10674 | 150857 | 4666 | 64291 |

\*COVID patient number has been enumerated using the government data displayed on the webpage https://ahmedabadcity.gov.in/portal/web?requestType=ApplicationRH&actionVal=loadCoronaRelatedDtls&queryType=Select&screenId=114 (Accessed on 14<sup>th</sup> June, 2020) https://www.covid19india.org/ (Accessed on 15<sup>th</sup> June, 2020)

For all the three genes, the abundance in the wastewater influent was higher in the samples of May 27, which was suggested by the larger CT values than in the sample of May 8. This result is consistent with the infection numbers in the area; In India, the daily new confirmed cases in the previous 10 days of the survey days were approximately double, 3000 and 6000 in the case of May 8 and May 27, respectively. A precise number of active cases for the city i.e. Ahmedabad and India, obtained by deducting recovered cases from total confirmed cases since 17 March, 2020 has been presented in Table 1. The consistency between abundance of SARS-CoV-2 genetic materials and number of confirmed cases was also observed in the previous reports in Australia, France, Italy, Spain and Japan (Ahmed et al 2020; Hata et al 2020; Randazzo et al 2020; Wurtzer et al 2020), showing that WBE is promising as a surveillance tool of COVID-19 spread in a community. In future studies, standards of SARS-CoV-2 RNA should be used for determining actual concentrations of SARS-CoV-2 RNA in wastewater samples India and for comparing results with other studies.

The final effluent samples taken on May 8 and May 27 were negative (CT values > 40) with all three SARS-CoV-2 genes examined, showing that the genes were significantly reduced by the UASB treatment and aeration pond. The SARS-CoV-2 genes were shown to decrease during treatments by a secondary treatment and a tertiary treatment by decantation, coagulation, flocculation, sand filtration, disinfection, NaClO, peracetic acid or UV in Spain and Italy (Randazzo et al 2020;Rimoldi et al 2020). To date, our results suggest the reduction of SARS-CoV-2 RNA after UASB treatment and aeration pond for the first time.

In this study, we employed PEG precipitation method for concentrating viruses, which have been used for detecting SARS-CoV-2 RNA (Hata et al 2020; La Rosa et al 2020; Wu et al. 2020). The recovery of spiked MS2 was stable in our study (CT values ranging from 22.2 to 22.46). In Hata et al 2020, the recovery of indigenous F-phages during PEG concentration was high and stable (57% in geometric mean), indicating an efficient recovery of SARS-CoV-2 with PEG concentration method. In the future, virus concentration performance of PEG method should be evaluated with multiple other virus concentration methods (e.g., ultrafiltration, aluminum hydroxide adsorption-precipitation) in analysing raw wastewater in India.

A comparative analyses of protocol and obtained results of genetic material detection of SARS-CoV-2 has been presented in Table 2. Overall, we have successfully detected ORF1ab, N protein genes and S protein genes, from Indian wastewater samples by RT-PCR and observed a significant decrease in the final effluents after UASB treatment and aeration pond. Ten times increase in the number of gene copies was noticed between 8 and 27 May, 2020 i.e. 0.78×10^2^ copies L^-1^ and 8.05×10^2^ copies L^-1^, respectively (Table 2), which corresponds to more than the doubling in the number of active COVID-19 patient in Ahmedabad city i.e. 4912 and 10674 individuals on 8 and 27 May, respectively (Table 1). Number of gene copies was found comparable to that reported in the untreated wastewaters of Australia, China and Turkey and lower than that of the USA, France and Spain.

**Table 2:**
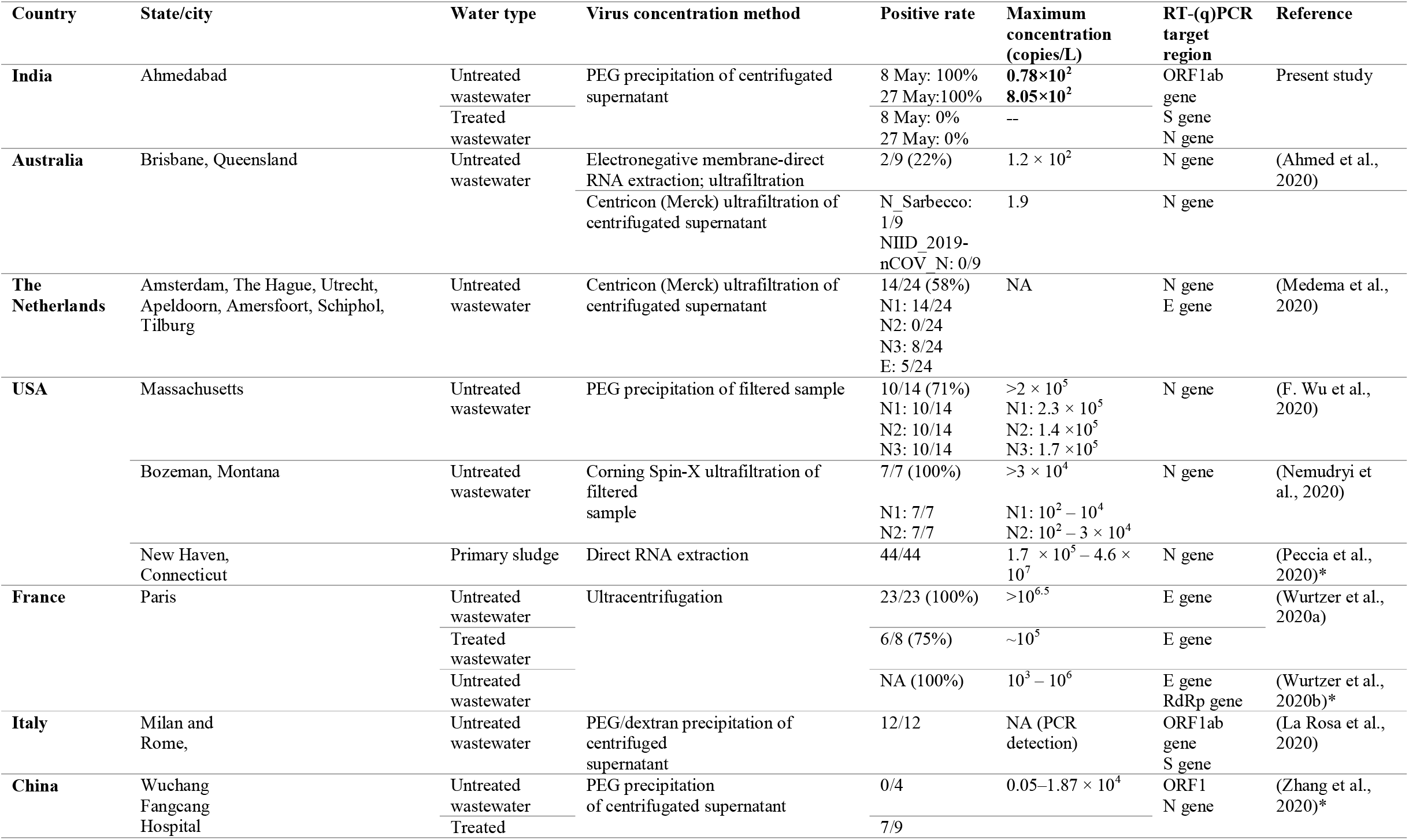
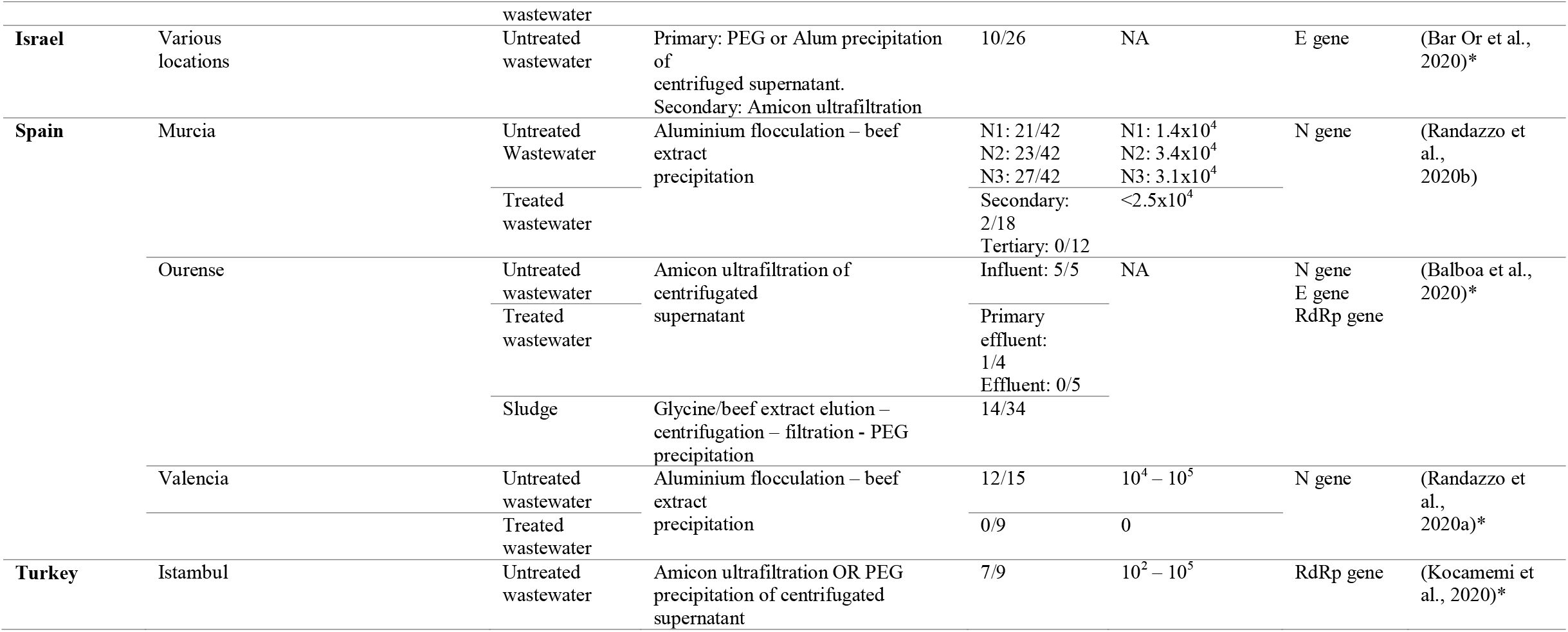
Comparative details of reported molecular detection of SARS-CoV-2 in the wastewater of various countries.

In summary, results proved the capability of wastewater based epidemiology in Indian settings and strongly advocates that despite the lack of quality sewer infrastructure or other wastewater collection issues, WBE can be unimaginably applicable and thus we strongly advocates the implementation environmental surveillances of CVOID-19 pandemic in India, starting with major cities.

## Conclusions

Broad estimation of COVID-19 prevalence at a community level in a populous country like India is posing a great difficulty yet essential for government and hospital officials responsible for pandemic crisis management of COVID-19. Owing to slow individual testing, money involved and impracticality of clinical testing of COVID-19 infected or suspected individuals, there needs an environmental surveillances of COID-19 like through detection of genetic material of SARS-CoV-2 in sewage. While the world is still requiring the proof of WBE concept, India needed a indigenous proof of concept and its applicability. In this context, we could achieve two major outcomes: i) for the first time ever in India and top 10 efforts in the world, we could isolate SARS-CoV-2 and detect its gene even during lockdown period with help of government organization i.e. GBRC; ii) temporal variation in Ct value proved the capability of WBE surveillance in India; and iii) for the third time in the world treated water has been analysed for the presence confirmation of genetic material of SARS-CoV-2. The results were of good resolution with significant indicative of temporal variation in COVID-19 patient loadings. Our result have effectively demonstrated that conventional treatment plant is capable of removing all the genetic materials of SARS.CoV-2. In the context that the asymptomatic people as well as those with mild symptoms are advised to stay at home the true scale of the pandemic must use the environmental surveillances of SARS-CoV-2 in wastewater to supplement the individual testing and timely identification. While in a country like India where sewer systems are not complete and only a part of waste is being received at WWTPs, it is essential to study after each treatment stage to indicate the effect of treatment. This will help curing the commonly perceived fear of the commons pertaining to the effectiveness of treatment plants as well as the transmission through wastewater.

## Data Availability

We will provide on the request basis.

## Notes

The authors declare no competing financial interest.

## Acknowledgement

We acknowledge the unimaginable support from every corner as naming all of them involved or helped during analyses or sampling or protocol development would be difficult. Special mentions are Prof. Sudhir Jain, IITGN, Dr. Sunita Varjani, GPCB, Ms. Nehal, Ahmedabad Municipal Corporation, and Dr. Neelam M Nathani, GBRC.

---

**Figure 1:**
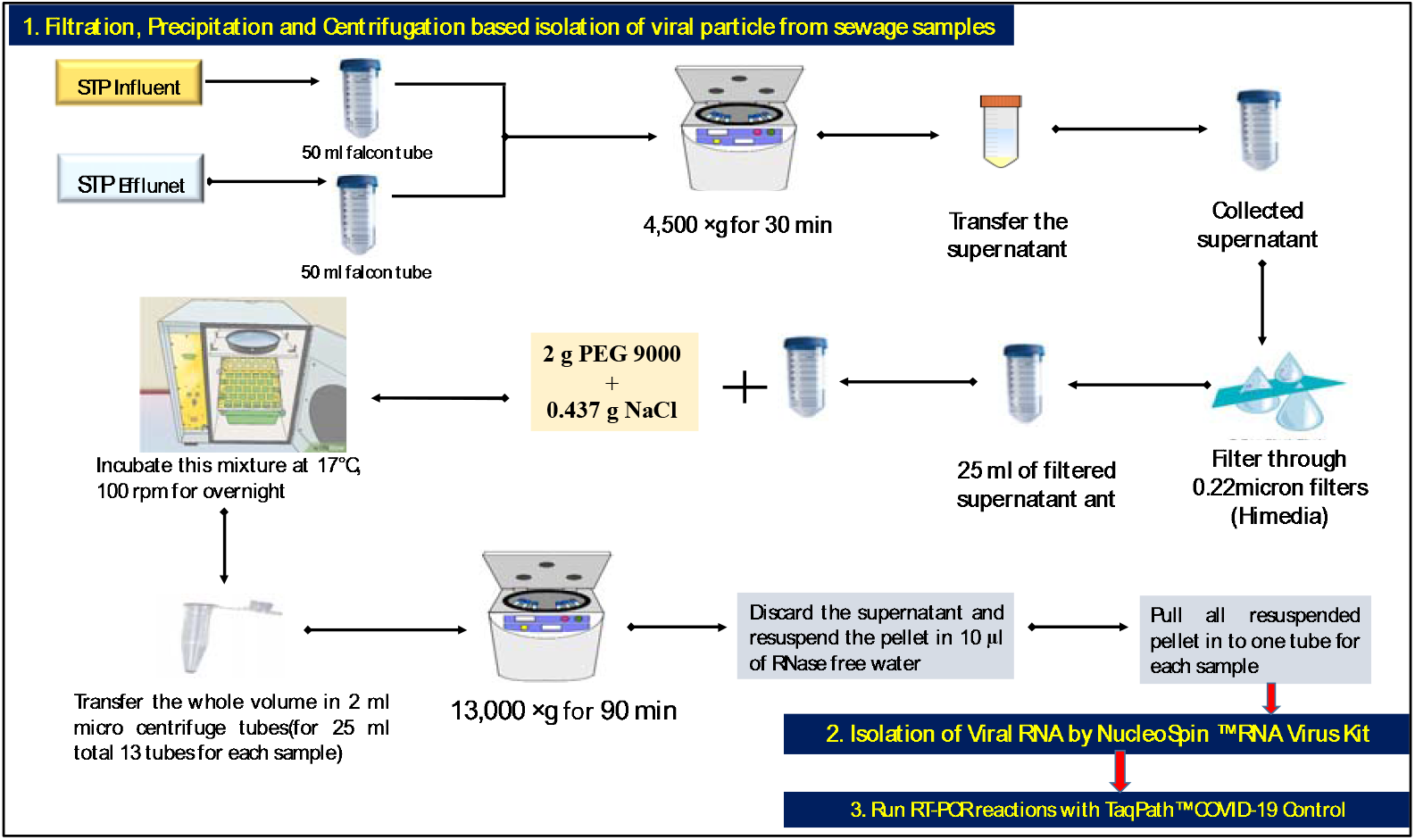
Illustrative flowchart of the modified polyethylene glycol (PEG) precipitation of centrifugated viral isolation from the wastewater samples followed by RNA isolation and reverse transcription polymerase chain reaction (RT-PCR) with 40 amplification cycles and final effluents.

**Figure 2:**
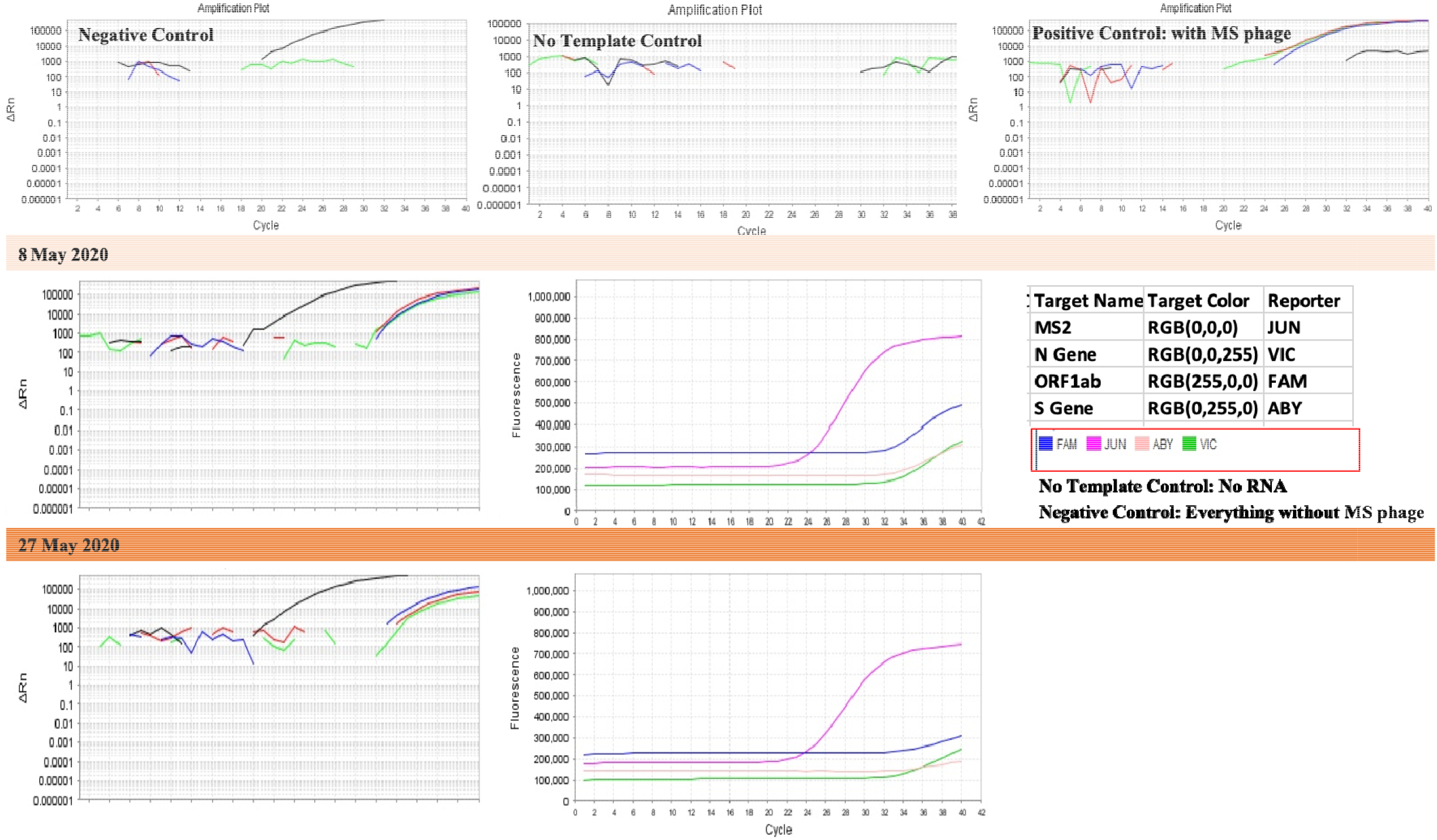
Amplification plots obtained through RT-PCR illustrating temporal variation through Ct value and Rn value between the samples of 8 and 27 May, 2020. Three control samples have also been provided.

